# No single biological phenotype exists in polycystic ovary syndrome: evidence from cross-space phenotyping

**DOI:** 10.64898/2026.07.09.26357636

**Authors:** Natalia Piórkowska, Alan Ostromęcki, Grzegorz Franik, Anna Bizoń

## Abstract

**Context:** Polyendocrine metabolic ovarian syndrome (PMOS), formerly known as polycystic ovary syndrome (PCOS), is a biologically heterogeneous disorder, yet previous clustering studies have reported inconsistent phenotype structures. Whether these discrepancies reflect methodological variability or genuine multidimensional disease biology remains unknown.

**Objective:** To determine whether independently derived endocrine, metabolic, inflammatory, and thyroid phenotypes represent the same underlying biological structure or capture distinct dimensions of PMOS heterogeneity.

**Design:** Cross-sectional observational study using a cross-space phenotyping framework.

**Setting:** Tertiary referral outpatient endocrinology and gynecology clinic.

**Participants:** A total of 1,286 women were diagnosed with PCOS according to the Rotterdam criteria.

**Methods:** Four predefined biological spaces (endocrine, metabolic, inflammatory, and thyroid) were analyzed independently. Within each space, standardized preprocessing, dimensionality reduction, and unsupervised clustering were performed. Cluster robustness was evaluated using bootstrap resampling, while agreement between independently derived phenotypes was quantified using the adjusted Rand index (ARI). Biological relevance was assessed using independent non-circular validation with variables excluded from phenotype derivation. Sensitivity analyses compared complete-case and imputed datasets.

**Results:** All four biological spaces produced highly stable clustering solutions (bootstrap ARI: endocrine 0.915, metabolic 0.964, inflammatory 0.930, thyroid 0.990). Despite this robustness, agreement between independently derived phenotypes remained consistently low. The highest concordance was observed between metabolic and inflammatory phenotypes (ARI = 0.208), followed by endocrine and metabolic phenotypes (ARI = 0.159), whereas agreement involving thyroid phenotypes was close to zero. Independent non-circular validation confirmed that all identified phenotypes represented biologically coherent patient subgroups beyond the variables used for clustering. Sensitivity analyses demonstrated high agreement between complete-case and imputed solutions, supporting the robustness of the findings.

**Conclusions:** Stable biological phenotypes exist within individual physiological domains of PMOS but do not converge into a single overarching biological phenotype. These findings support a multidimensional model of PMOS heterogeneity in which endocrine, metabolic, inflammatory, and thyroid systems describe complementary rather than interchangeable aspects of disease biology. Cross-space phenotyping provides a general framework for investigating biological heterogeneity in complex disorders and may facilitate future precision medicine approaches.

## 1. Introduction

Polycystic ovary syndrome (PCOS) affects an estimated 4–21% of women of reproductive age worldwide, depending on the diagnostic criteria applied [1]. The prevalence of PCOS also varies considerably across European countries and regions [2].

PCOS is a complex endocrine and metabolic disorder characterized by hyperandrogenism, ovulatory dysfunction, and polycystic ovarian morphology [3].

Beyond reproductive manifestations, PCOS is associated with a broad spectrum of metabolic [4], cardiovascular [5], and psychological and metabolic comorbidities [6], including obesity [7], insulin resistance, type 2 diabetes [8], dyslipidemia, endometrial disorders [9], depression, obstructive sleep apnea, and anxiety [10].

For many years, researchers and clinicians have emphasized that the term “polycystic ovary syndrome” does not adequately reflect the complex and heterogeneous nature of the disorder, given its wide range of reproductive, endocrine, metabolic, and clinical manifestations and associated comorbidities [11]. This complexity is reflected in the substantial heterogeneity of the affected population, which comprises distinct phenotypic subgroups that may differ in their clinical manifestations, underlying pathophysiology, and therapeutic needs, highlighting the potential value of more individualized approaches to patient management [12]. Furthermore, the EGOI-PCOS perspective conceptualizes PCOS as a complex endocrine–metabolic condition characterized by marked interindividual variation across multiple biological domains, including reproductive, metabolic, inflammatory, and endocrine processes. Consistent with this broader view of disease heterogeneity, the 2023 International Evidence-Based Guideline for the Assessment and Management of Polycystic Ovary Syndrome advocates a comprehensive, patient-centered assessment of metabolic, cardiovascular, and endocrine risk profiles extending beyond classification based solely on conventional diagnostic phenotypes [13]. Reflecting this growing recognition of the complex and multidimensional nature of the disorder, the recent global consensus published by Teede et al. recommended renaming PCOS as polyendocrine metabolic ovarian syndrome (PMOS) [14].

The conventional phenotypic classification of PCOS is based on the Rotterdam criteria and distinguishes four phenotypes according to different combinations of hyperandrogenism, ovulatory dysfunction, and polycystic ovarian morphology [15]. Although clinically useful, this classification captures only selected manifestations of the disorder and may not fully reflect its broader biological heterogeneity. Increasing evidence suggests that women with PCOS exhibit distinct reproductive and metabolic profiles and trajectories that extend beyond conventional diagnostic phenotypes [16,17].

Therefore, data-driven approaches, including unsupervised clustering, have increasingly been applied to identify biologically distinct patient subgroups.

However, previous clustering studies have reported inconsistent phenotype structures, potentially reflecting differences in study populations, variable selection, analytical methods, or the multidimensional nature of the disorder itself.

A critical unresolved question is whether endocrine, metabolic, inflammatory, and thyroid abnormalities in PMOS converge into a common underlying biological structure or represent distinct and complementary dimensions of heterogeneity.

Previous clustering studies have typically derived phenotypes from combined sets of clinical and biochemical variables, implicitly assuming that a single overarching phenotype structure can adequately capture the biological complexity of PMOS. Whether independently derived phenotypes across different physiological domains identify the same individuals or instead reveal discordant biological architectures remains unknown.

Given the biological heterogeneity of PMOS and the inconsistent findings of previous clustering studies, we aimed to determine whether independently derived endocrine, metabolic, inflammatory, and thyroid phenotypes reflect a shared biological architecture or represent distinct dimensions of PMOS heterogeneity.

## 2. Methods

### 2.1. Study design and objectives

This retrospective cross-sectional study was designed to determine whether PCOS can be represented by a single latent biological phenotype or whether different physiological systems define distinct and only partially overlapping patterns of patient heterogeneity.

Rather than deriving a single clustering solution from all available variables, we implemented a cross-space phenotyping framework in which biologically related variables were grouped *a priori* into four independent biological spaces representing endocrine, metabolic, inflammatory, and thyroid physiology. Each biological space was analyzed separately using an identical unsupervised machine learning pipeline.

The primary objective was to quantify the agreement between independently derived biological phenotypes across these four physiological domains. Specifically, we evaluated whether women assigned to the same phenotype within one biological space remained grouped together when phenotypes were derived from another biological space.

Secondary objectives were to (i) evaluate the internal stability and reproducibility of phenotype identification within each biological space, (ii) characterize the resulting biological phenotypes using clinical and laboratory variables, (iii) assess phenotype generalizability using independent biological validation, and (iv) determine the robustness of the analytical framework through predefined sensitivity analyses evaluating the impact of missing data handling and alternative analytical assumptions.

The study was designed to investigate biological concordance rather than diagnostic classification or prediction. Accordingly, no supervised learning models were developed, and all analyses focused exclusively on unsupervised phenotype discovery and quantitative comparison of independently derived biological representations of PCOS.

### 2.2. Study population

The study included women diagnosed with PCOS who were evaluated at a tertiary referral outpatient endocrinology and gynecology clinic between 2018 and 2025. PCOS was diagnosed according to the Rotterdam criteria, requiring the presence of at least two of the following features after exclusion of alternative etiologies: (i) oligo- or anovulation, (ii) clinical and/or biochemical hyperandrogenism, and (iii) polycystic ovarian morphology on ultrasonography [15].

Women with incomplete diagnostic evaluation precluding confirmation of the PCOS diagnosis were excluded. Additional exclusion criteria were applied according to the routine clinical diagnostic protocol used to exclude alternative causes of hyperandrogenism and ovulatory dysfunction before establishing the diagnosis of PCOS.

The analysis was restricted to women with PCOS. Healthy controls were not included because the objective of the study was to investigate biological heterogeneity within the PCOS population rather than to develop a diagnostic classification model.

Because laboratory investigations were performed according to routine clinical practice, availability of individual laboratory measurements differed between patients. Consequently, the number of participants included in each biological space varied according to the availability of variables required for that specific analysis.

The study was conducted in accordance with the Declaration of Helsinki and was approved by the Bioethical Committee of Wroclaw Medical University (approval no. 254/2021). All data were analyzed in anonymized form.

### 2.3. Clinical and laboratory measurements

Clinical, anthropometric, and laboratory data were retrospectively extracted from the institutional electronic medical records and laboratory information system. All measurements were obtained as part of routine diagnostic evaluation of women with PCOS.

The endocrine domain included variables describing hypothalamic–pituitary–ovarian and adrenal function, including luteinizing hormone (LH), follicle stimulating hormone (FSH), the LH/FSH ratio, anti-Müllerian hormone (AMH), total and free testosterone, sex hormone binding globulin (SHBG), free androgen index (FAI), dehydroepiandrosterone sulfate (DHEAS), and morning and evening cortisol measurements where available.

The metabolic domain comprised fasting and 2-hour oral glucose tolerance test (OGTT) glucose and insulin measurements, lipid profile components (total cholesterol, low-density lipoprotein cholesterol (LDL-C), high-density lipoprotein cholesterol (HDL-C), and triglycerides (TG)), and routinely derived metabolic indices including homeostatic model assessment of insulin resistance (HOMA-IR), quantitative insulin sensitivity check index (QUICKI), triglyceride–glucose index (TyG), TG/HDL-C ratio, and non-HDL-C.

The inflammatory domain included complete blood count parameters, leukocyte differential measurements, C-reactive protein (CRP), and derived inflammatory indices including neutrophil-to-lymphocyte ratio (NLR), platelet-to-lymphocyte ratio (PLR), lymphocyte-to-monocyte ratio (LMR), and systemic immune-inflammation index (SII).

The thyroid domain included thyroid-stimulating hormone (TSH), free thyroxine (fT4), and thyroid peroxidase antibodies (anti-TPO).

Laboratory analyses were performed in the institutional certified clinical laboratory. Variable transformations and derived indices were generated automatically within the predefined computational pipeline before downstream analyses.

### 2.4. Definition of biological spaces

The central premise of this study was that PCOS represents a multidimensional biological disorder rather than a condition characterized by a single latent phenotype. Accordingly, phenotypes were not derived from all available variables simultaneously. Instead, clinically related variables were grouped *a priori* into four independent biological spaces representing distinct physiological systems involved in PCOS pathophysiology.

The composition of each biological space was defined before any unsupervised analyses based on current knowledge of PCOS biology and routine clinical interpretation, rather than statistical associations observed within the study cohort.

The **endocrine space** comprised variables reflecting hypothalamic–pituitary–ovarian and adrenal function, including LH, FSH, the LH/FSH ratio, total testosterone, SHBG, AMH, and DHEAS, where available.

The **metabolic space** included variables describing glucose homeostasis, insulin sensitivity, and lipid metabolism, including fasting and post-load glucose, insulin concentrations, lipid profile components, HOMA-IR, QUICKI, TyG, TG/HDL-C ratio, and non-HDL-C.

The **inflammatory space** comprised complete blood count parameters, leukocyte differential measurements, CRP, and derived hematological inflammatory indices.

The **thyroid space** included markers of thyroid function and autoimmunity, including TSH, ffT4, anti-TPO antibodies, and additional thyroid-related laboratory variables available in the study cohort.

Each biological space was analyzed independently throughout the analytical workflow. Variables were not shared across spaces during dimensionality reduction or clustering, ensuring that each clustering solution represented an independent biological representation of the same patient cohort.

Variables used for phenotype derivation were distinguished from those reserved for subsequent phenotype characterization and independent biological validation, thereby minimizing circular interpretation.

### 2.5. Data harmonization and preprocessing

Data preprocessing was performed using a predefined computational pipeline established before any unsupervised analyses. The objective was to generate a harmonized analytical dataset while minimizing investigator-dependent decisions.

Clinical and laboratory variables were extracted from the institutional database and harmonized using predefined mapping rules. Laboratory measurements representing the same biological parameter but recorded under different names or abbreviations were consolidated into standardized feature identifiers.

Quality control included verification of variable identity, assessment of measurement consistency, detection of implausible values, evaluation of variable completeness, and exclusion of variables with ambiguous biological interpretation. No variables were selected or excluded on the basis of clustering performance or downstream analytical results.

Continuous variables were inspected for distributional characteristics, and predefined transformations were applied where appropriate. Derived clinical indices were calculated before dimensionality reduction according to standard clinical definitions.

To ensure comparability across variables measured on different numerical scales, continuous variables were standardized independently within each biological space before dimensionality reduction and clustering.

The preprocessing pipeline was fully reproducible. Intermediate datasets generated at each analytical stage were saved automatically and served as the exclusive input for subsequent analyses, ensuring complete traceability of the computational workflow.

### 2.6. Missing data handling

Missing laboratory measurements were expected because not all participants underwent identical diagnostic evaluations during routine clinical care.

The primary analyses were performed using complete-case datasets generated independently for each biological space. As a predefined sensitivity analysis, complementary datasets were generated after univariate imputation of missing values using the SimpleImputer implementation from scikit-learn. Imputation was performed independently within each biological space, preventing information sharing between physiological domains.

The influence of missing data handling was evaluated by comparing clustering solutions obtained from complete-case and imputed datasets using the adjusted Rand index (ARI). Imputed datasets were used exclusively for sensitivity analyses and were not considered replacements for the primary complete-case analyses.

### 2.7. Dimension reduction

Dimensionality reduction was performed independently within each predefined biological space using principal component analysis (PCA). Continuous variables were standardized before PCA, and separate PCA models were estimated for each biological space.

The number of retained principal components was determined using a predefined cumulative explained variance threshold of 80%, applied uniformly across all biological spaces. Principal component scores were subsequently used as input for clustering analyses.

PCA served exclusively as a computational step to reduce redundancy among correlated laboratory variables and was not used for biological interpretation of individual features.

### 2.8. Unsupervised clustering

Unsupervised clustering was performed independently within each biological space using Gaussian Mixture Models (GMMs) fitted to the corresponding principal component representations.

Candidate models with different numbers of clusters were evaluated using Bayesian Information Criterion (BIC), silhouette coefficient, bootstrap-derived cluster stability, posterior assignment probabilities, and predefined minimum cluster-size requirements. Bootstrap reproducibility was assessed using 100 resampling iterations, and agreement between bootstrap-derived and original clustering solutions was quantified using the ARI.

The primary clustering solution for each biological space was selected according to the predefined model-selection framework rather than any single optimization criterion. All clustering parameters and selection rules were specified before biological interpretation, and no clustering solutions were modified after inspection of downstream results.

### 2.9. Cross-space agreement analysis

The primary objective of the study was to quantify the extent to which independently derived biological phenotypes identified the same individuals across distinct physiological domains. After clustering had been completed separately within each biological space, agreement between clustering solutions was evaluated without modifying or aligning the original cluster assignments.

Pairwise comparisons were performed between all biological spaces, resulting in six independent cross-space comparisons: endocrine–metabolic, endocrine–inflammatory, endocrine–thyroid, metabolic–inflammatory, metabolic–thyroid, and inflammatory–thyroid. Each comparison assessed whether patients grouped together within one biological space remained grouped together when phenotypes were derived from another physiological domain.

Agreement between clustering solutions was quantified using the ARI, which measures similarity between two independent partitions while correcting for agreement expected by chance. ARI values close to one indicate highly concordant phenotype assignments, whereas values close to zero indicate agreement no greater than expected by random partitioning. Negative values indicate disagreement exceeding chance expectation.

To determine whether the observed agreement exceeded random expectation, statistical significance was evaluated using permutation testing. Cluster labels from one biological space were randomly permuted while preserving cluster sizes, and the adjusted Rand index was recalculated repeatedly to generate an empirical null distribution. Observed ARI values were subsequently compared with this null distribution to obtain permutation-based *P* values.

Cross-space agreement analyses were performed using the primary clustering solutions and repeated as part of predefined sensitivity analyses, including complete-case and imputed datasets. This strategy enabled assessment of whether biological concordance remained stable under alternative missing-data assumptions.

Importantly, no attempt was made to maximize agreement between biological spaces by harmonizing feature sets, modifying cluster assignments, or selecting alternative clustering solutions after inspection of cross-space results. Each biological space was analyzed independently throughout the computational workflow, and agreement metrics were calculated only after all clustering procedures had been finalized.

The purpose of the cross-space agreement analysis was not to identify a single optimal clustering solution but to determine whether different physiological systems describe a common latent biological phenotype or instead capture complementary and partially independent dimensions of PCOS heterogeneity. Consequently, cross-space agreement constituted the primary endpoint of the study.

These findings directly support the primary study hypothesis that distinct physiological systems capture complementary rather than interchangeable dimensions of PCOS biology.

### 2.10. Biological characterization

Following phenotype identification, each cluster was characterized using the original clinical and laboratory variables to facilitate biological interpretation of the independently derived endocrine, metabolic, inflammatory, and thyroid phenotypes. This stage was performed only after completion of all clustering procedures and did not influence phenotype definition, model selection, or cross-space agreement analyses.

Within each biological space, descriptive summaries were generated separately for all identified phenotypes. Continuous variables were summarized using appropriate measures of central tendency and dispersion according to their distribution, whereas categorical variables were presented as counts and percentages. Differences between phenotypes were assessed using statistical methods appropriate for the distribution and measurement scale of each variable. To control for multiple testing, *P* values were adjusted using the Benjamini–Hochberg false discovery rate (FDR) procedure where applicable.

Biological characterization was intended to describe the clinical and biochemical profiles associated with each phenotype rather than to provide evidence supporting the validity of the clustering procedure. Accordingly, interpretation focused on coherent biological patterns across related variables instead of isolated statistically significant findings. Cluster-specific profiles were interpreted within the physiological context of the corresponding biological space.

Because some characterization variables also contributed to phenotype derivation, these analyses were considered descriptive. Independent evaluation of phenotype generalizability using complementary variables that were not involved in phenotype construction was performed separately, as described in the following section.

The biological coherence of these phenotypes was subsequently evaluated using independent validation variables that were not involved in phenotype derivation.

### 2.11. Independent Biological Validation

To determine whether the identified phenotypes reflected biologically meaningful patient stratification rather than intrinsic properties of the variables used for clustering, an independent biological validation strategy was implemented. Validation analyses were designed to evaluate phenotype differences using clinical and laboratory variables that had not been used to construct the corresponding biological space whenever possible.

For each biological space, variables directly involved in phenotype derivation were distinguished from independent validation variables before any statistical analyses were performed. Consequently, validation focused on assessing whether cluster membership was associated with complementary biological characteristics rather than reproducing differences that were inherently encoded in the clustering procedure.

Independent validation analyses were performed separately for each biological space using predefined validation datasets generated within the computational pipeline. Continuous validation variables were compared across phenotypes using appropriate parametric or nonparametric statistical tests according to data distribution, whereas categorical variables were analyzed using contingency table methods. False discovery rate correction using the Benjamini–Hochberg procedure was applied to account for multiple comparisons.

The primary objective of this validation stage was not to maximize statistical significance but to determine whether independently derived biological phenotypes demonstrated coherent differences in complementary physiological domains. Validation was therefore interpreted as evidence supporting the biological plausibility and external consistency of the identified phenotypes rather than as confirmation of the clustering algorithm itself.

When complete independence between clustering variables and validation variables was not biologically feasible because of the close physiological relationships within a given domain, interpretation emphasized complementary biological information rather than variables directly determining cluster assignment. This approach minimized circular interpretation while preserving clinically meaningful characterization of the identified phenotypes.

Independent biological validation was conducted after all clustering procedures, model selection, and cross-space agreement analyses had been finalized. Validation results did not influence phenotype definitions, cluster selection, or any other stage of the analytical workflow, thereby maintaining strict separation between phenotype discovery and phenotype evaluation.

### 2.12. Sensitivity analyses

A series of predefined sensitivity analyses was performed to evaluate the robustness of the identified biological phenotypes and the primary cross-space agreement findings. These analyses were designed to determine whether the principal conclusions remained consistent under alternative analytical assumptions and were specified before interpretation of the biological results.

First, the influence of missing data handling was assessed by comparing clustering solutions obtained from complete-case datasets with those generated after imputation. Agreement between the corresponding clustering solutions was quantified using the ARI independently for each biological space. This analysis evaluated whether phenotype identification was substantially affected by the chosen missing-data strategy.

Second, cross-space agreement analyses were repeated using clustering solutions derived from both complete-case and imputed datasets. This approach allowed assessment of whether the observed concordance or discordance between biological spaces represented a stable biological finding rather than an artifact of data completeness.

Third, additional diagnostic analyses were performed for biological spaces exhibiting lower clustering robustness or potentially unstable partition structures. These analyses included evaluation of cluster size distributions, posterior assignment probabilities, bootstrap reproducibility, and model-selection diagnostics to determine whether individual clustering solutions met predefined quality criteria.

Finally, biological characterization and independent validation analyses were interpreted jointly with the corresponding clustering stability metrics. Phenotypes demonstrating high reproducibility across bootstrap resampling and consistent behavior across sensitivity analyses were considered robust. Conversely, clustering solutions showing reduced stability or evidence of dependence on analytical assumptions were interpreted cautiously and regarded as exploratory.

Sensitivity analyses were not used to modify the primary clustering solutions or redefine biological phenotypes. Instead, they provided an independent assessment of the reliability and reproducibility of the analytical framework and supported transparent interpretation of the study findings.

### 2.13. Statistical analysis

Statistical analyses were conducted according to a predefined analysis plan established before interpretation of the clustering results.

Continuous variables were summarized as mean ± standard deviation (SD) or median (interquartile range [IQR]), as appropriate, whereas categorical variables were presented as counts and percentages. Comparisons between phenotypes were performed using parametric or nonparametric methods according to variable distribution and measurement scale. Categorical variables were compared using contingency table analyses. Where applicable, *P* values were adjusted using the Benjamini–Hochberg false discovery rate procedure.

The primary study endpoint was cross-space agreement between independently derived biological phenotypes, quantified using the ARI. Statistical significance of cross-space agreement was evaluated using permutation testing, whereas phenotype robustness was assessed using bootstrap resampling. Agreement between complete-case and imputed clustering solutions was evaluated as part of the predefined sensitivity analyses.

All statistical tests were two-sided, and *P* <0.05 was considered statistically significant unless otherwise specified following multiple-testing correction. Effect sizes and 95% confidence intervals were reported where appropriate.

### 2.14. Software and reproducibility

All computational analyses were performed in Python using a fully reproducible analytical pipeline. The workflow consisted of sequential, self-contained computational notebooks implementing predefined analytical stages, including data harmonization, preprocessing, dimensionality reduction, clustering, cross-space agreement analysis, biological characterization, independent biological validation, sensitivity analyses, and final quality control.

The analytical pipeline was deterministic, with the output of each stage serving as the exclusive input for the subsequent stage. Intermediate datasets, model objects, summary tables, quality-control reports, and graphical outputs were generated automatically, ensuring complete traceability of the analytical workflow. Randomized procedures were performed using predefined random seeds to maximize computational reproducibility.

Analyses were performed in Python using NumPy, pandas, SciPy, scikit-learn, statsmodels, Matplotlib, and related scientific computing libraries. Exact package versions, including the Python interpreter version, are provided in the accompanying GitHub repository in the computational environment file generated with the analytical pipeline.

The complete computational pipeline, including executable notebooks, metadata, and analytical outputs, will be made publicly available through a version-controlled GitHub repository upon publication to enable independent reproduction of all analyses presented in this study.

## 3. Results

### 3.1. Cohort Characteristics

The study included women with PCOS who fulfilled the Rotterdam diagnostic criteria and underwent routine clinical and laboratory evaluation at a tertiary referral outpatient clinic. Demographic characteristics together with the principal endocrine, metabolic, inflammatory, and thyroid measurements are summarized in Table 1.

**Table 1.**
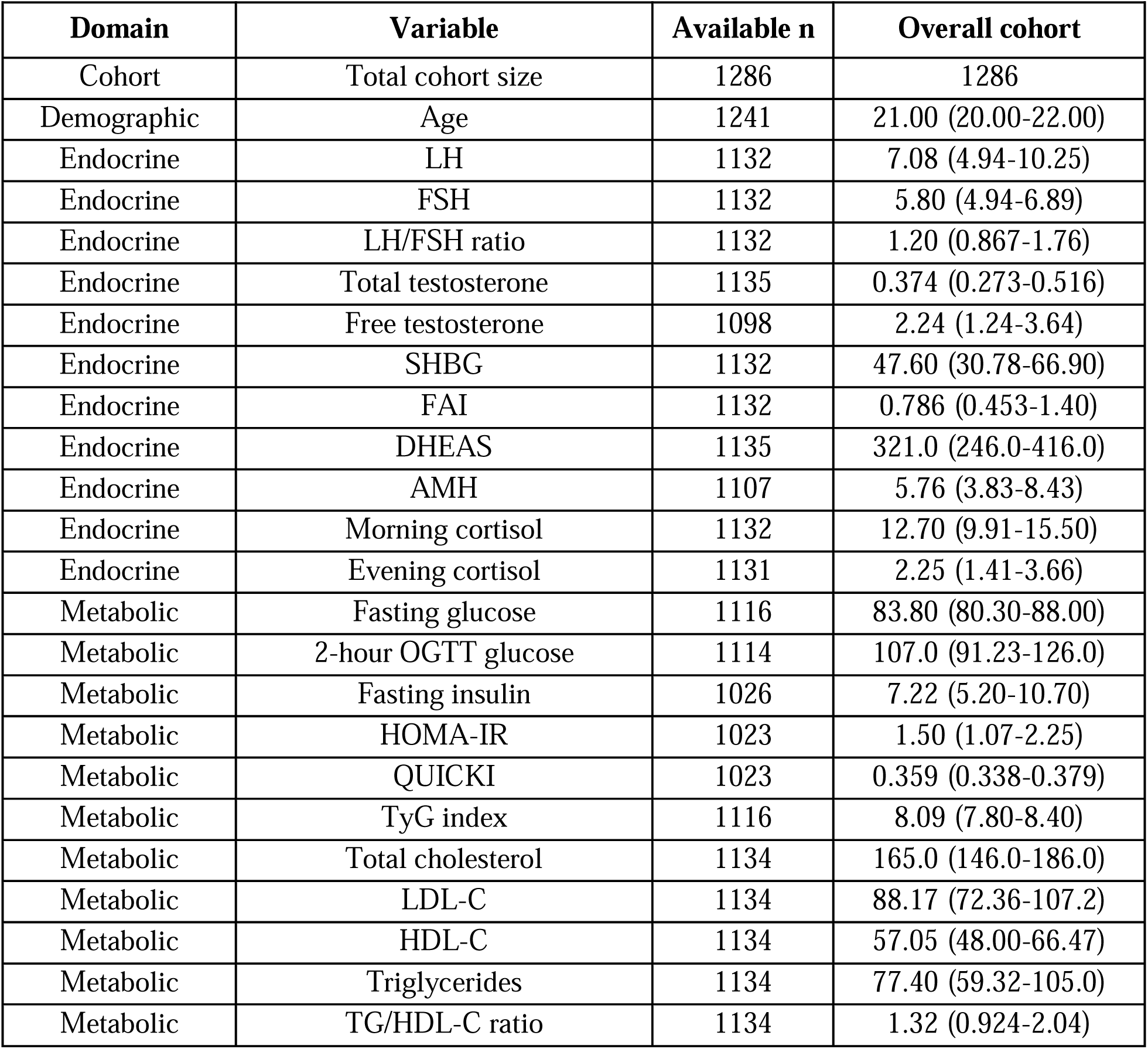
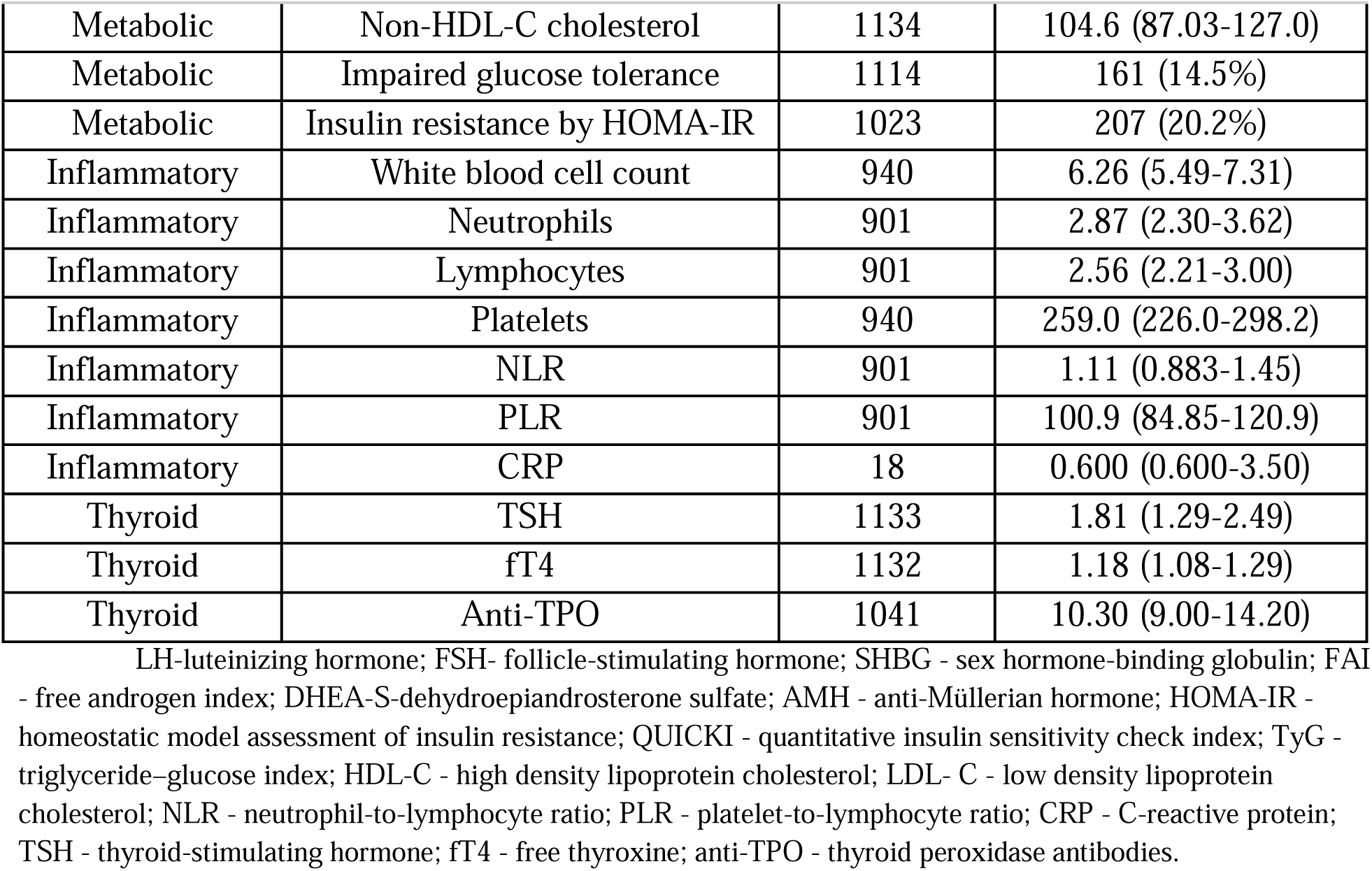
Baseline demographic, clinical, and laboratory characteristics of the study cohort.

| Domain | Variable | Available n | Overall cohort |
| --- | --- | --- | --- |
| Cohort | Total cohort size | 1286 | 1286 |
| Demographic | Age | 1241 | 21.00 (20.00-22.00) |
| Endocrine | LH | 1132 | 7.08 (4.94-10.25) |
| Endocrine | FSH | 1132 | 5.80 (4.94-6.89) |
| Endocrine | LH/FSH ratio | 1132 | 1.20 (0.867-1.76) |
| Endocrine | Total testosterone | 1135 | 0.374 (0.273-0.516) |
| Endocrine | Free testosterone | 1098 | 2.24 (1.24-3.64) |
| Endocrine | SHBG | 1132 | 47.60 (30.78-66.90) |
| Endocrine | FAI | 1132 | 0.786 (0.453-1.40) |
| Endocrine | DHEAS | 1135 | 321.0 (246.0-416.0) |
| Endocrine | AMH | 1107 | 5.76 (3.83-8.43) |
| Endocrine | Morning cortisol | 1132 | 12.70 (9.91-15.50) |
| Endocrine | Evening cortisol | 1131 | 2.25 (1.41-3.66) |
| Metabolic | Fasting glucose | 1116 | 83.80 (80.30-88.00) |
| Metabolic | 2-hour OGTT glucose | 1114 | 107.0 (91.23-126.0) |
| Metabolic | Fasting insulin | 1026 | 7.22 (5.20-10.70) |
| Metabolic | HOMA-IR | 1023 | 1.50 (1.07-2.25) |
| Metabolic | QUICKI | 1023 | 0.359 (0.338-0.379) |
| Metabolic | TyG index | 1116 | 8.09 (7.80-8.40) |
| Metabolic | Total cholesterol | 1134 | 165.0 (146.0-186.0) |
| Metabolic | LDL-C | 1134 | 88.17 (72.36-107.2) |
| Metabolic | HDL-C | 1134 | 57.05 (48.00-66.47) |
| Metabolic | Triglycerides | 1134 | 77.40 (59.32-105.0) |
| Metabolic | TG/HDL-C ratio | 1134 | 1.32 (0.924-2.04) |
| Metabolic | Non-HDL-C cholesterol | 1134 | 104.6 (87.03-127.0) |
| Metabolic | Impaired glucose tolerance | 1114 | 161 (14.5%) |
| Metabolic | Insulin resistance by HOMA-IR | 1023 | 207 (20.2%) |
| Inflammatory | White blood cell count | 940 | 6.26 (5.49-7.31) |
| Inflammatory | Neutrophils | 901 | 2.87 (2.30-3.62) |
| Inflammatory | Lymphocytes | 901 | 2.56 (2.21-3.00) |
| Inflammatory | Platelets | 940 | 259.0 (226.0-298.2) |
| Inflammatory | NLR | 901 | 1.11 (0.883-1.45) |
| Inflammatory | PLR | 901 | 100.9 (84.85-120.9) |
| Inflammatory | CRP | 18 | 0.600 (0.600-3.50) |
| Thyroid | TSH | 1133 | 1.81 (1.29-2.49) |
| Thyroid | ft4 | 1132 | 1.18 (1.08-1.29) |
| Thyroid | Anti-TPO | 1041 | 10.30 (9.00-14.20) |

Because laboratory investigations were performed according to routine clinical practice, the availability of individual variables differed across participants. Consequently, the number of women included in subsequent analyses varied between biological spaces depending on data completeness. This analytical strategy was predefined and enabled independent evaluation of endocrine, metabolic, inflammatory, and thyroid phenotypes without introducing information leakage between biological domains.

### 3.2. Identification of Biological Phenotypes

Independent unsupervised clustering was performed within each predefined biological space to identify endocrine, metabolic, inflammatory, and thyroid phenotypes. The stability of the resulting clustering solutions was evaluated using bootstrap resampling, with agreement between the original and bootstrap-derived cluster assignments quantified by the ARI.

The endocrine, metabolic, and inflammatory spaces demonstrated consistently high clustering stability, with bootstrap ARI values of 0.915, 0.964, and 0.930, respectively (Figure 1). These findings indicate that the identified phenotypes were highly reproducible across repeated resampling and were unlikely to represent unstable data partitions.

**Figure 1.**
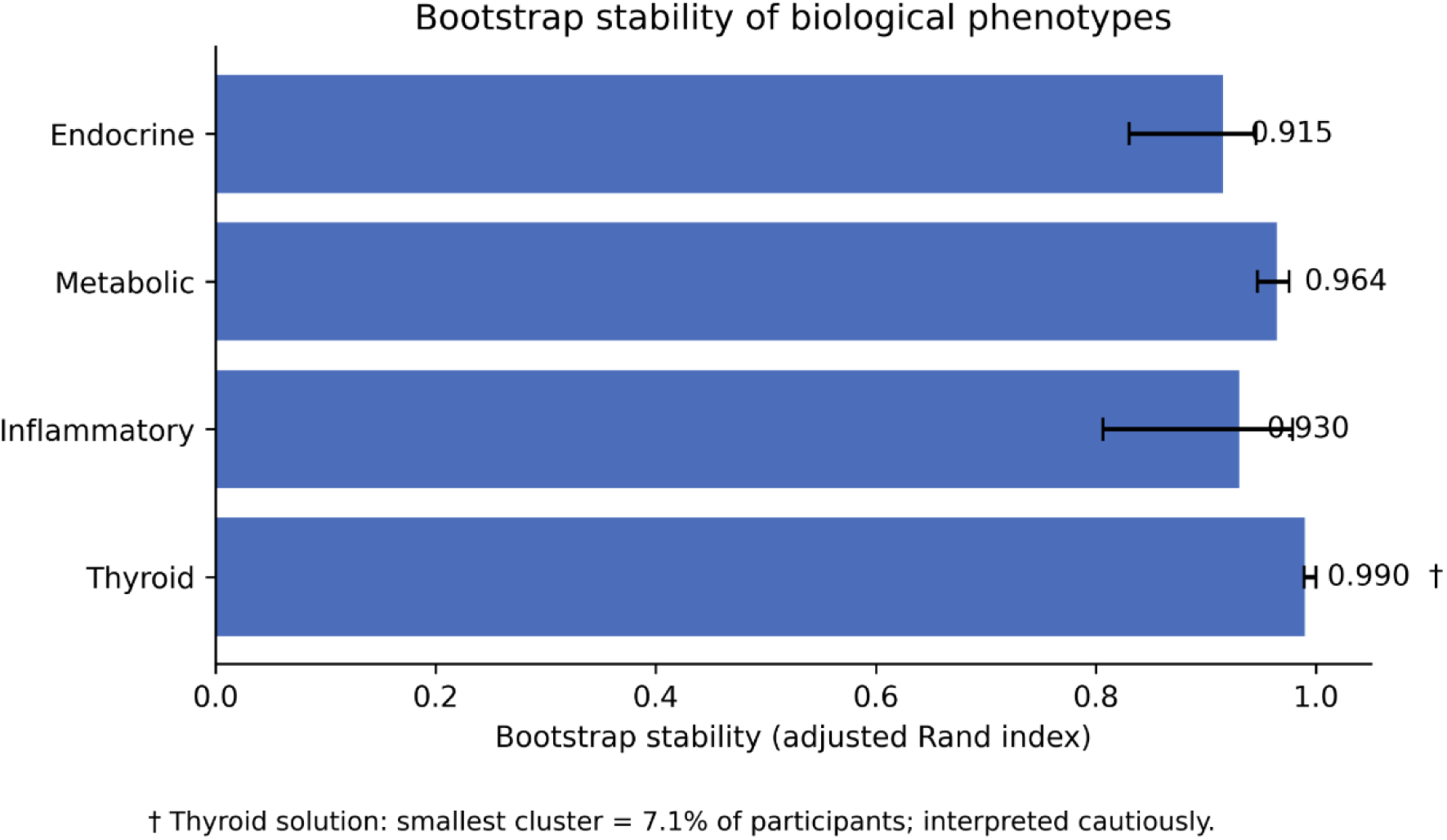
Bootstrap stability of independently derived biological phenotypes across the four predefined biological spaces.

The thyroid space demonstrated the highest numerical stability (bootstrap ARI = 0.990). However, the selected clustering solution contained a small cluster representing approximately 7% of the study population. Although this solution satisfied the predefined model-selection criteria, the thyroid phenotypes were interpreted cautiously in subsequent analyses because of the limited size of this subgroup.

Overall, all four biological spaces produced reproducible clustering solutions that met the predefined stability criteria and were therefore retained for subsequent cross-space agreement analyses, biological characterization, and independent biological validation.

### 3.3. Cross-Space Agreement

After phenotype identification within each biological space, we quantified whether independently derived endocrine, metabolic, inflammatory, and thyroid phenotypes classified the same women into corresponding subgroups. Cross-space concordance was generally low, indicating that the biological spaces captured distinct and only partially overlapping dimensions of PCOS heterogeneity.

The strongest agreement was observed between the metabolic and inflammatory spaces (ARI = 0.208), followed by endocrine and metabolic phenotypes (ARI = 0.159). These values indicate partial but limited concordance between metabolic dysregulation, inflammatory status, and endocrine phenotype structure (Figure 2).

**Figure 2.**
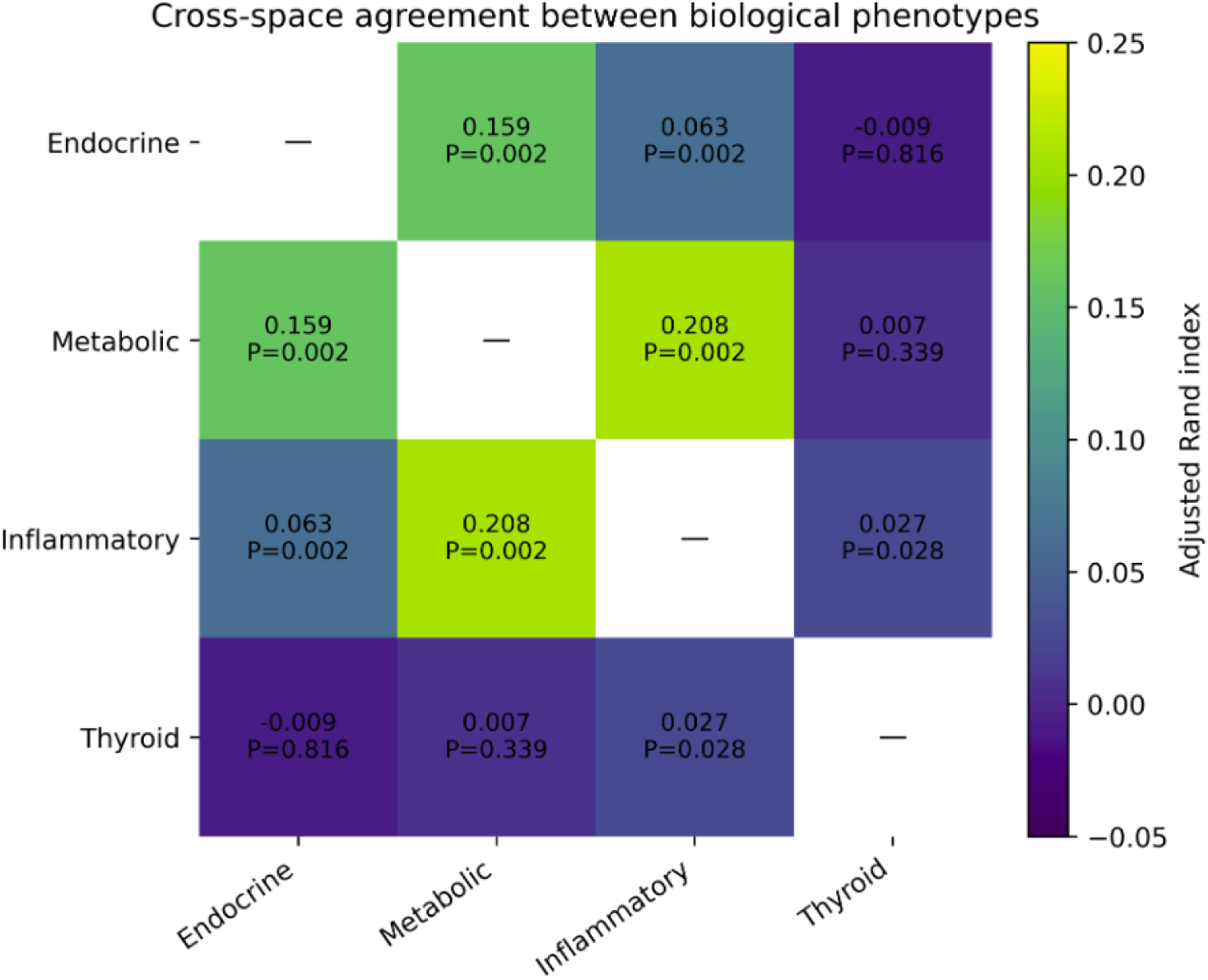
Cross-space agreement between independently derived biological phenotypes. Pairwise agreement between endocrine, metabolic, inflammatory, and thyroid phenotype assignments was quantified using the ARI. Higher ARI values indicate greater concordance between biological spaces. Values close to zero indicate little or no agreement beyond chance.

In contrast, agreement involving the thyroid space was close to zero, including endocrine–thyroid and metabolic–thyroid comparisons. This suggests that thyroid-defined phenotype identified a largely distinct subgroup structure that did not overlap meaningfully with endocrin or metabolic phenotypes.

Overall, the low cross-space agreement supports the central hypothesis that PCOS cannot be adequately represented by a single biological phenotype. Instead, endocrine, metabolic, inflammatory, and thyroid domains appear to organize patient heterogeneity in different and only partially concordant ways.

### 3.4. Biological Characterization of Identified Phenotypes

Following independent phenotype identification within each biological space, the resulting clusters were characterized using the original clinical and laboratory variables to facilitate biological interpretation. Distinct biological profiles were observed within all four physiological domains, demonstrating that the identified phenotypes reflected coherent patterns of biological variation rather than isolated differences in individual laboratory measurements.

Within the endocrine space, phenotypes were primarily differentiated by coordinated variation in reproductive hormone concentrations and androgen-related parameters, indicating distinct endocrine profiles among women with PCOS. Similarly, metabolic phenotypes differed in glucose homeostasis, insulin resistance, and lipid metabolism, supporting the presence of biologically meaningful metabolic heterogeneity within the cohort (Figure 3).

**Figure 3.**
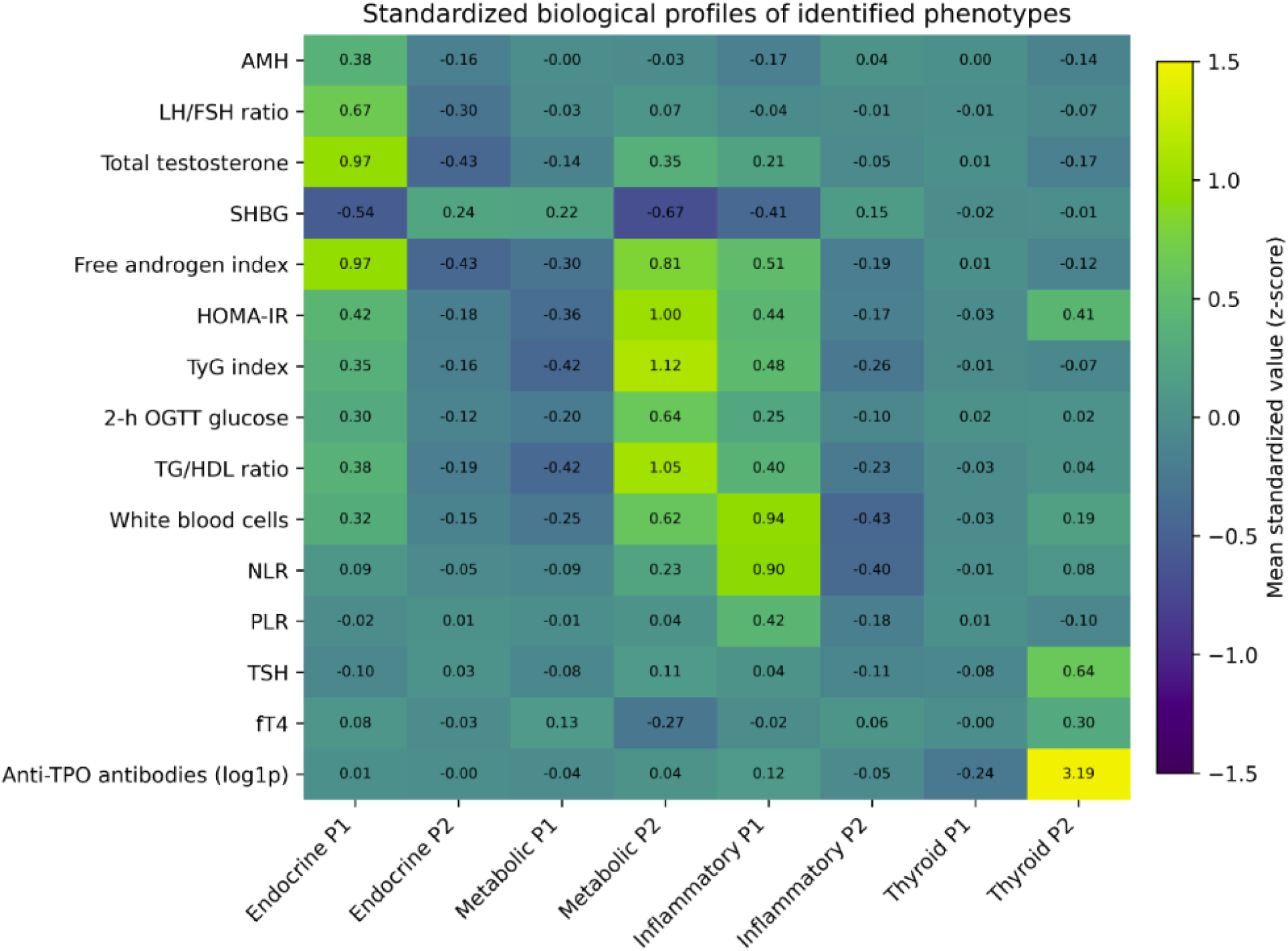
Standardized biological profiles of the endocrine, metabolic, inflammatory, and thyroid phenotypes. **Legend:** AMH - anti-Müllerian hormone; LH-luteinizing hormone; FSH - follicle-stimulating hormone; SHBG - sex hormone-binding globulin; HOMA-IR - homeostatic model assessment of insulin resistance; TyG - triglyceride–glucose index; OGTT - oral glucose tolerance test; TG - triglycerides; HDL-C - high density lipoprotei cholesterol; NLR - neutrophil-to-lymphocyte ratio; PLR - platelet-to-lymphocyte ratio; TSH - thyroid-stimulating hormone; fT4 - free thyroxine; anti-TPO - thyroid peroxidase antibodies.

Inflammatory phenotypes were characterized by consistent differences in leukocyte-derived inflammatory markers and systemic inflammatory status. Although these phenotypes were identified independently of the endocrine and metabolic spaces, they demonstrated internally coherent inflammatory profiles, indicating that inflammatory variation represents an additional dimension of PCOS heterogeneity (Table 2).

**Table 2.**
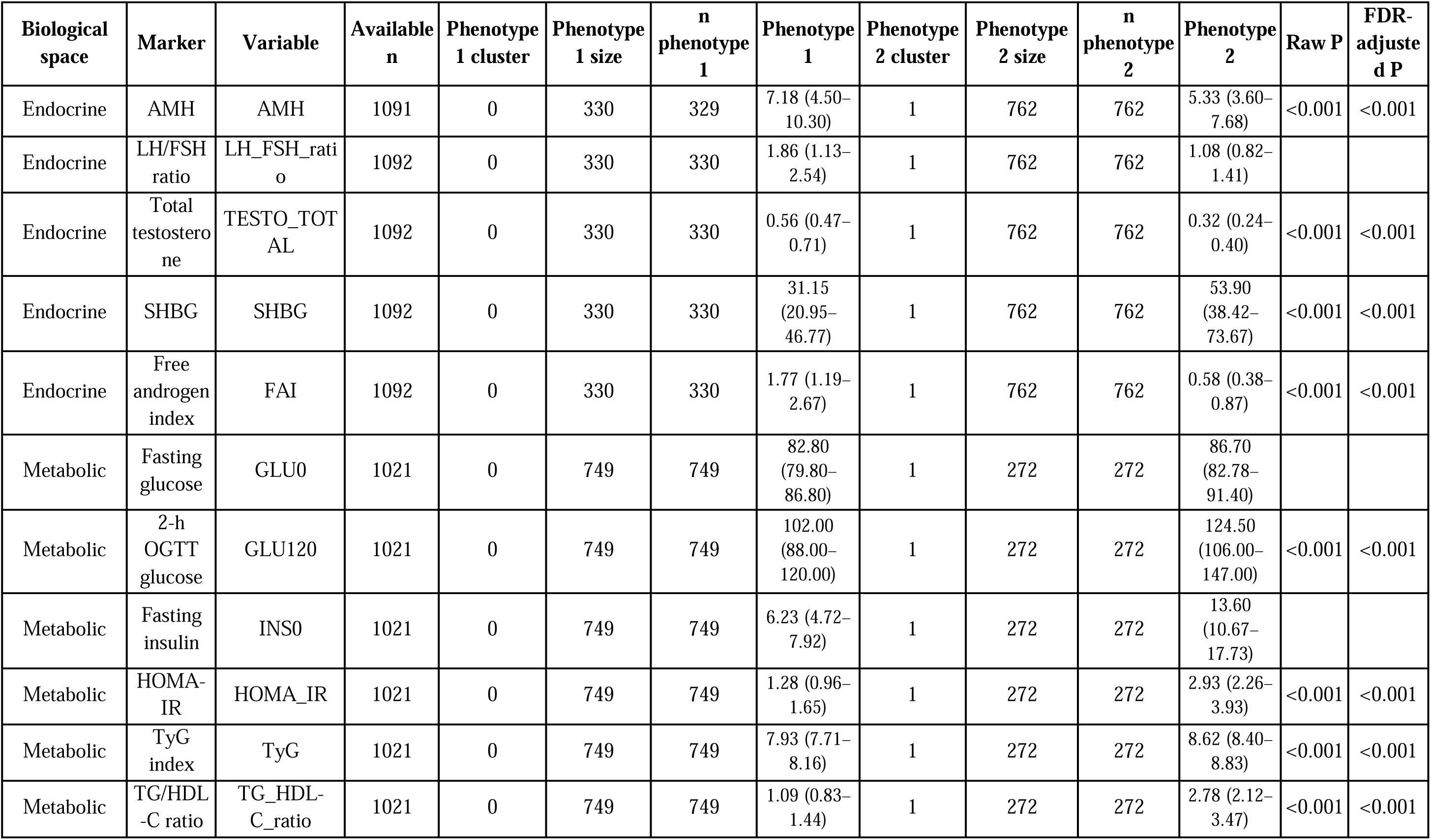
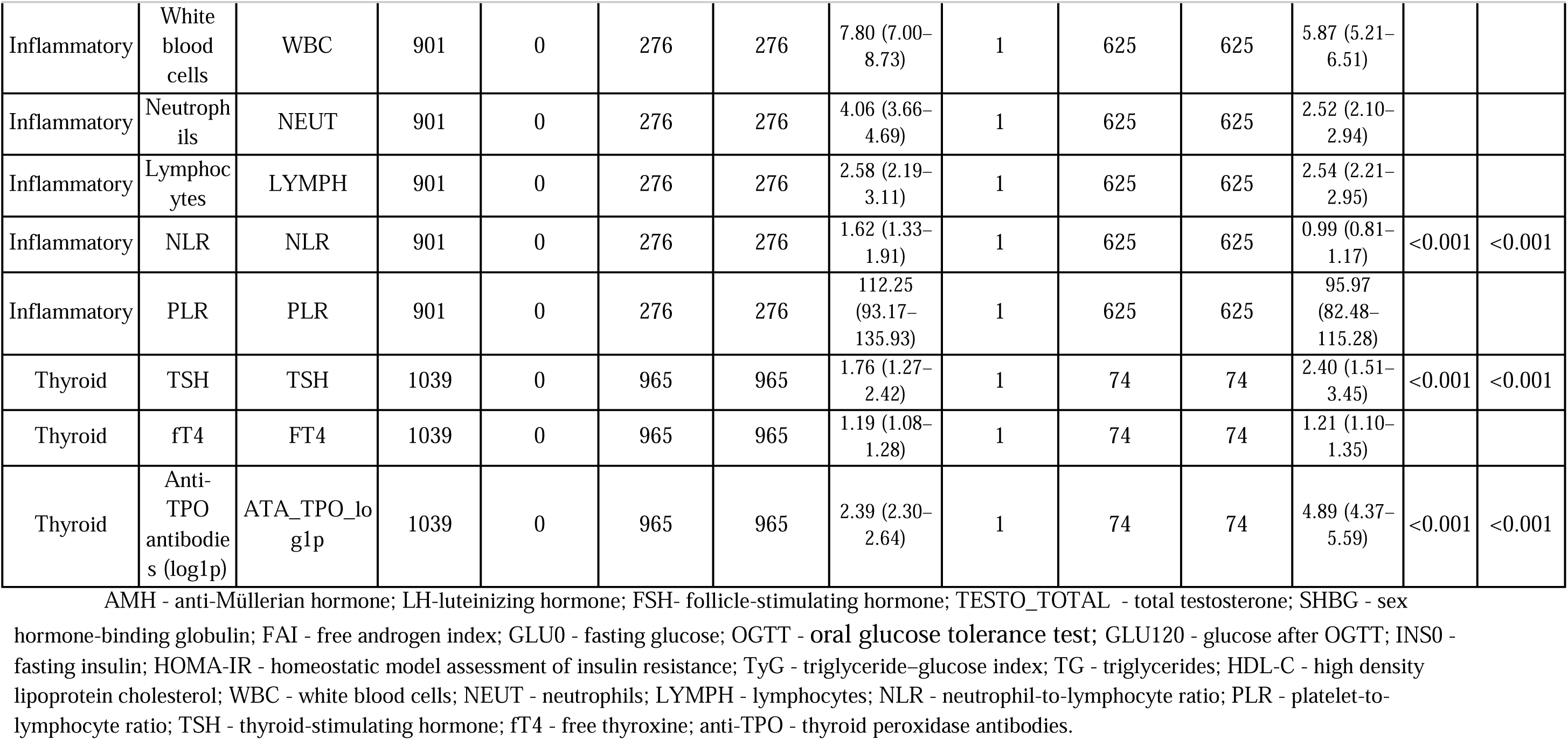
Biological characteristics of the endocrine, metabolic, inflammatory, and thyroid phenotypes identified by cross-space phenotyping.

| Biological space | Marker | Variable | Available n | Phenotype 1 cluster | Phenotype 1 size | n phenotype 1 | Phenotype 1 | Phenotype 2 cluster | Phenotype 2 size | n phenotype 2 | Phenotype 2 | Raw P | FDR-adjusted P |
| --- | --- | --- | --- | --- | --- | --- | --- | --- | --- | --- | --- | --- | --- |
| Endocrine | AMH | AMH | 1091 | 0 | 330 | 329 | 7.18 (4.50–10.30) | 1 | 762 | 762 | 5.33 (3.60–7.68) | <0.001 | <0.001 |
| Endocrine | LH/FSH ratio | LH_FSH_ratio | 1092 | 0 | 330 | 330 | 1.86 (1.13–2.54) | 1 | 762 | 762 | 1.08 (0.82–1.41) |  |  |
| Endocrine | Total testosterone | TESTO_TOTAL | 1092 | 0 | 330 | 330 | 0.56 (0.47–0.71) | 1 | 762 | 762 | 0.32 (0.24–0.40) | <0.001 | <0.001 |
| Endocrine | SHBG | SHBG | 1092 | 0 | 330 | 330 | 31.15 (20.95–46.77) | 1 | 762 | 762 | 53.90 (38.42–73.67) | <0.001 | <0.001 |
| Endocrine | Free androgen index | FAI | 1092 | 0 | 330 | 330 | 1.77 (1.19–2.67) | 1 | 762 | 762 | 0.58 (0.38–0.87) | <0.001 | <0.001 |
| Metabolic | Fasting glucose | GLU0 | 1021 | 0 | 749 | 749 | 82.80 (79.80–86.80) | 1 | 272 | 272 | 86.70 (82.78–91.40) |  |  |
| Metabolic | 2-h OGTT glucose | GLU120 | 1021 | 0 | 749 | 749 | 102.00 (88.00–120.00) | 1 | 272 | 272 | 124.50 (106.00–147.00) | <0.001 | <0.001 |
| Metabolic | Fasting insulin | INS0 | 1021 | 0 | 749 | 749 | 6.23 (4.72–7.92) | 1 | 272 | 272 | 13.60 (10.67–17.73) |  |  |
| Metabolic | HOMA-IR | HOMA_IR | 1021 | 0 | 749 | 749 | 1.28 (0.96–1.65) | 1 | 272 | 272 | 2.93 (2.26–3.93) | <0.001 | <0.001 |
| Metabolic | TyG index | TyG | 1021 | 0 | 749 | 749 | 7.93 (7.71–8.16) | 1 | 272 | 272 | 8.62 (8.40–8.83) | <0.001 | <0.001 |
| Metabolic | TG/HDL-C ratio | TG_HDL-C_ratio | 1021 | 0 | 749 | 749 | 1.09 (0.83–1.44) | 1 | 272 | 272 | 2.78 (2.12–3.47) | <0.001 | <0.001 |
| Inflammatory | White blood cells | WBC | 901 | 0 | 276 | 276 | 7.80 (7.00–8.73) | 1 | 625 | 625 | 5.87 (5.21–6.51) |  |  |
| Inflammatory | Neutrophils | NEUT | 901 | 0 | 276 | 276 | 4.06 (3.66–4.69) | 1 | 625 | 625 | 2.52 (2.10–2.94) |  |  |
| Inflammatory | Lymphocytes | LYMPH | 901 | 0 | 276 | 276 | 2.58 (2.19–3.11) | 1 | 625 | 625 | 2.54 (2.21–2.95) |  |  |
| Inflammatory | NLR | NLR | 901 | 0 | 276 | 276 | 1.62 (1.33–1.91) | 1 | 625 | 625 | 0.99 (0.81–1.17) | <0.001 | <0.001 |
| Inflammatory | PLR | PLR | 901 | 0 | 276 | 276 | 112.25 (93.17–135.93) | 1 | 625 | 625 | 95.97 (82.48–115.28) |  |  |
| Thyroid | TSH | TSH | 1039 | 0 | 965 | 965 | 1.76 (1.27–2.42) | 1 | 74 | 74 | 2.40 (1.51–3.45) | <0.001 | <0.001 |
| Thyroid | ft4 | FT4 | 1039 | 0 | 965 | 965 | 1.19 (1.08–1.28) | 1 | 74 | 74 | 1.21 (1.10–1.35) |  |  |
| Thyroid | Anti-TPO antibodies (log1p) | ATA_TPO_log1p | 1039 | 0 | 965 | 965 | 2.39 (2.30–2.64) | 1 | 74 | 74 | 4.89 (4.37–5.59) | <0.001 | <0.001 |

The thyroid space identified phenotypes differing in thyroid function and thyroid autoimmunity markers. Despite the high numerical stability of the clustering solution, interpretation of thyroid phenotypes was performed cautiously because one subgroup represented a relatively small proportion of the cohort (Table S1).

Taken together, these findings demonstrate that each biological space identified internally consistent physiological phenotypes while emphasizing different aspects of PCOS biology. The observed phenotype structures therefore support the concept that PCOS heterogeneity is multidimensional rather than driven by a single dominant biological process.

### 3.5. Independent Biological Validation

To evaluate whether the identified phenotypes represented biologically meaningful patient subgroups rather than mathematical partitions of the data, we performed an independent non-circular validation using variables that were not involved in phenotype derivation.

Across all four biological spaces, independently selected validation variables demonstrated significant differences between the identified phenotypes (Table 3). Endocrine phenotypes exhibited distinct metabolic characteristics, whereas metabolic phenotypes differed in endocrine markers that had not contributed to clustering. Similarly, inflammatory phenotypes demonstrated significant variation in metabolic and endocrine measures, while thyroid phenotypes showed differences in complementary biological characteristics beyond thyroid-specific variables.

**Table 3.** Independent non-circular biological validation of the identified endocrine, metabolic, inflammatory, and thyroid phenotypes.

| Biological space | Independent validation marker | Variable | Statistical test | n | Statistic | Raw P | FDR-adjusted P | Significant after FDR (0.05) |
| --- | --- | --- | --- | --- | --- | --- | --- | --- |
| Endocrine | AMH | AMH | Kruskal | 1091 | 53.111 | <0.001 | <0.001 | Yes |
| Endocrine | HOMA-IR | HOMA_IR | Kruskal | 1010 | 87.437 | <0.001 | <0.001 | Yes |
| Endocrine | TyG index | TyG | Kruskal | 1077 | 56.631 | <0.001 | <0.001 | Yes |
| Endocrine | TG/HDL-C ratio | TG_HDL-C_ratio | Kruskal | 1092 | 75.469 | <0.001 | <0.001 | Yes |
| Endocrine | 2-h OGTT glucose | GLU120 | Kruskal | 1075 | 30.32 | <0.001 | <0.001 | Yes |
| Endocrine | NLR | NLR | Kruskal | 888 | 10.871 | <0.001 | 0.002 | Yes |
| Metabolic | Free androgen index | FAI | Kruskal | 1021 | 225.872 | <0.001 | <0.001 | Yes |
| Metabolic | Total testosterone | TESTO_TOTAL | Kruskal | 1021 | 43.853 | <0.001 | <0.001 | Yes |
| Metabolic | SHBG | SHBG | Kruskal | 1021 | 248.071 | <0.001 | <0.001 | Yes |
| Metabolic | TSH | TSH | Kruskal | 1021 | 5.845 | 0.016 | 0.028 | Yes |
| Metabolic | NLR | NLR | Kruskal | 886 | 40.138 | <0.001 | <0.001 | Yes |
| Inflammatory | AMH | AMH | Kruskal | 900 | 8.995 | 0.003 | 0.005 | Yes |
| Inflammatory | HOMA-IR | HOMA_IR | Kruskal | 887 | 108.938 | <0.001 | <0.001 | Yes |
| Inflammatory | TyG index | TyG | Kruskal | 888 | 98.679 | <0.001 | <0.001 | Yes |
| Inflammatory | TG/HDL-C ratio | TG_HDL-C_ratio | Kruskal | 900 | 94.076 | <0.001 | <0.001 | Yes |
| Inflammatory | 2-h OGTT glucose | GLU120 | Kruskal | 887 | 21.897 | <0.001 | <0.001 | Yes |
| Inflammatory | Free androgen index | FAI | Kruskal | 900 | 69.704 | <0.001 | <0.001 | Yes |
| Inflammatory | Total testosterone | TESTO_TOTAL | Kruskal | 901 | 10.545 | 0.001 | 0.002 | Yes |
| Inflammatory | SHBG | SHBG | Kruskal | 900 | 88.448 | <0.001 | <0.001 | Yes |

The consistency of these cross-domain associations indicates that the identified phenotypes captured broader biological organization rather than variation restricted to the variables used for clustering. Importantly, these findings were obtained using independent validation variables, thereby avoiding circular interpretation.

Overall, the non-circular validation analyses provide independent biological support for the identified phenotypes and demonstrate that the observed subgroup structure reflects biologically coherent patterns extending beyond the original feature space used for phenotype identification. (Table S2)

### 3.6. Sensitivity Analyses

Sensitivity analyses assessed whether the identified biological phenotypes were robust to the handling of missing data. Clustering solutions obtained from complete-case datasets were compared with corresponding solutions derived after within-space imputation using the ARI.

Agreement between complete-case and imputed solutions was high across biological spaces: endocrine ARI = 0.872, metabolic ARI = 0.915, inflammatory ARI = 0.737, and thyroid ARI = 0.992. These findings indicate that phenotype identification was not primarily driven by the chosen missing-data strategy.

Cross-space agreement patterns remained consistent after imputation, supporting the robustness of the primary conclusion that endocrine, metabolic, inflammatory, and thyroid spaces describe only partially overlapping dimensions of PCOS heterogeneity.

### 3.7. Independent Biological Validation

To evaluate whether the identified phenotypes represented biologically meaningful patient subgroups rather than mathematical partitions of the data, we performed an independent non-circular validation using variables that were not involved in phenotype derivation.

Across all four biological spaces, independently selected validation variables demonstrated significant differences between the identified phenotypes (Table 3). Endocrine phenotypes exhibited distinct metabolic characteristics, whereas metabolic phenotypes differed in endocrine markers that had not contributed to clustering. Similarly, inflammatory phenotypes demonstrated significant variation in metabolic and endocrine measures, while thyroid phenotypes showed differences in complementary biological characteristics beyond thyroid-specific variables.

The consistency of these cross-domain associations indicates that the identified phenotypes captured broader biological organization rather than variation restricted to the variables used for clustering. Importantly, these findings were obtained using independent validation variables, thereby avoiding circular interpretation.

Overall, the non-circular validation analyses provide independent biological support for the identified phenotypes and demonstrate that the observed subgroup structure reflects biologically coherent patterns extending beyond the original feature space used for phenotype identification.

## 4. Discussion

### 4.1. Principal findings

PCOS is widely described as a heterogeneous disorder, yet almost all previous attempts to resolve this heterogeneity have relied on a single clustering solution derived from a single, pooled feature space. Implicit in that approach is the assumption that one biological partition can adequately summarize a woman’s endocrine, metabolic, inflammatory, and thyroid status at the same time. The present study tested this assumption directly, using a cross-space phenotyping framework that derived phenotypes independently within each biological domain before asking whether they described the same underlying patient structure.

Our principal finding is that this assumption does not hold. Endocrine, metabolic, and inflammatory phenotypes were each highly stable and internally coherent, and the thyroid space showed the highest numerical stability of all four domains, yet agreement between spaces was consistently low, and was weakest for the thyroid space. Because these phenotypes were additionally supported by non-circular biological validation using variables that had not contributed to clustering, this discordance cannot be attributed to overfitting or unstable partitions of the data. Instead, the four biological spaces appear to encode genuinely different, only partially overlapping, axes of patient heterogeneity.

This distinguishes the present work from prior PCOS clustering research, which has generally asked which single clustering solution best represents the syndrome. By instead quantifying agreement between independently derived, domain-specific phenotypes, we show that the search for one optimal solution may itself be misconceived: several biologically valid partitions of the same population can coexist, each emphasizing a different physiological system and therefore each answering a different biological question.

Rather than identifying a single optimal biological phenotype, our findings demonstrate that PCOS comprises multiple complementary physiological architectures that coexist within the same patient population.

### 4.2. Relation to previous clustering studies

That PCOS clustering studies rarely agree with one another is not a new observation; what has been missing is a framework that explains why. Placing our findings alongside three influential PCOS clustering studies, each of which used a different combination of variables, illustrates how cross-space phenotyping can reconcile this apparent inconsistency rather than simply restate it.

Dapas et al. performed hierarchical clustering in a genotyped discovery cohort of 893 women with PCOS using a comparatively narrow panel of reproductive and metabolic quantitative traits [18]. They identified two reproducible subtypes, a “reproductive” subtype with higher LH and SHBG and lower BMI and insulin, and a “metabolic” subtype with higher BMI, glucose, and insulin and lower SHBG and LH, and showed that these subtypes tracked with distinct common genetic loci. This two-cluster solution is broadly consistent with our endocrine and metabolic spaces considered separately: LH-, SHBG-, and androgen-driven variation on one hand, and glucose-, insulin-, and lipid-driven variation on the other. Critically, however, our cross-space agreement analysis shows that these two axes are only weakly correlated within the same cohort (endocrine-metabolic ARI = 0.159). This suggests that the reproductive-metabolic dichotomy described by Dapas et al. may reflect the two dominant, only partly overlapping, gradients that emerge whenever reproductive-endocrine and metabolic variables are combined into a single feature space, rather than a single unified reproductive-metabolic axis of disease.

Gao et al. recently analyzed 11,908 women across five international cohorts using a broader panel of nine clinical variables spanning anthropometric, reproductive, and endocrine domains, including AMH, and identified four reproducible subtypes, hyperandrogenic, obesity-predominant, high-SHBG, and high-LH-AMH, each with distinct longitudinal reproductive and metabolic trajectories [16]. The larger number of clusters relative to Dapas et al. is consistent with our findings: adding further endocrine dimensions and anthropometric information to a combined feature space increases the number of only weakly correlated axes of variation that a single clustering solution must accommodate, and each additional axis has the potential to fragment the resulting partition further. Notably, although our endocrine space did not include AMH or anthropometric variables as clustering inputs, post hoc validation showed that AMH nonetheless differed strongly across endocrine phenotypes, consistent with AMH marking a partially distinct sub-dimension of endocrine biology that is capable of shaping cluster structure when included.

Other studies illustrate the same principle using yet other variable combinations. Van der Ham et al., analyzing 2,502 women in a study also published in this journal, clustered on a panel that added androstenedione, DHEAS, cortisol, and antral follicle count to lipid and blood pressure variables, and obtained three subtypes, metabolic, reproductive, and an undifferentiated “background” group [19]. Elsayed et al. described PCOS phenotypes using a still different clinical variable combination [17]. Each of these studies used a defensible, biologically motivated variable set; none is more “correct” than the others.

Considered together, we propose the following synthesis: previous clustering studies were probably each correct in describing the data they analyzed, because they were, in effect, sampling different combinations of only partially overlapping biological spaces. When reproductive-endocrine, metabolic, anthropometric, and occasionally thyroid or immune variables are pooled into a single feature space before clustering, the resulting partition is highly sensitive to exactly which variables are included and how strongly each domain is represented, precisely because, as our cross-space agreement analysis demonstrates directly, these domains are only weakly concordant within the same women. Two, three, or four clusters can all be legitimate descriptions of the same underlying cohort, differing not because of methodological error but because they represent different projections of a genuinely multidimensional biological structure. This reframing is, to our knowledge, the principal conceptual contribution of the present study: rather than adding another clustering solution to an already crowded literature, we provide direct, quantitative evidence for why that literature has been unable to converge, and a framework, cross-space phenotyping, for interpreting future studies within it.

### 4.3. Biological interpretation

Rather than reviewing the individual clusters within each biological space in turn, it is more informative to ask why four physiological systems, measured in the same women, would be expected to generate only partially overlapping subgroup structures in the first place.

The endocrine space is organized primarily around hypothalamic-pituitary-ovarian axis activity, including gonadotropin secretion, ovarian androgen output, and SHBG-mediated androgen bioavailability. This axis is governed by neuroendocrine feedback loops that are only loosely coupled to peripheral energy metabolism, so endocrine phenotype structure need not track a woman’s metabolic status.

The metabolic space, by contrast, is dominated by insulin resistance and its downstream effects on glucose handling and lipid metabolism. Insulin resistance can arise from adiposity, genetic susceptibility, or intrinsic post-receptor signaling defects that are largely independent of ovarian steroidogenesis. Although insulin can stimulate ovarian androgen production, this interaction is neither obligatory nor uniform across women with PCOS, which limits the extent to which metabolic and endocrine phenotype structures can be expected to coincide.

The inflammatory space captures chronic low-grade inflammation, reflected in leukocyte-derived indices such as the neutrophil-to-lymphocyte ratio. Low-grade inflammation is mechanistically linked to insulin resistance and adiposity, which plausibly explains why the inflammatory and metabolic spaces showed the strongest, albeit still partial, agreement of any pair of spaces in our analysis. However, inflammatory activation can also be driven by factors unrelated to metabolic status, such as subclinical infection or unmeasured lifestyle exposures, which limits complete concordance between these two domains.

The thyroid space, finally, is shaped substantially by autoimmune thyroid disease, whose prevalence is increased in PCOS but whose pathogenesis, rooted in immune dysregulation and thyroid peroxidase autoantibody production, is largely distinct from the endocrine and metabolic pathways that define the syndrome’s cardinal features. This mechanistic separateness is consistent with the near-zero cross-space agreement observed for the thyroid space in our analysis.

Taken together, these four axes correspond to only partially overlapping regulatory networks: the hypothalamic-pituitary-ovarian axis, insulin-glucose signaling, the innate immune and inflammatory response, and thyroid autoimmunity are interconnected at the level of whole-body physiology, but each is governed by substantially independent regulatory machinery. Discordance between biological spaces is therefore not a statistical artifact but the expected consequence of profiling a syndrome whose cardinal features arise from multiple, only loosely coupled, physiological systems.

### 4.4. Implications for precision medicine

Why should this matter to a clinician managing a woman with PCOS? Because it implies that no single biological phenotype, and no single biomarker, can be expected to fully describe an individual patient’s risk profile.

Current practice often implicitly assumes that a patient’s PCOS presentation can be captured by a small number of features, such as degree of hyperandrogenism or presence of obesity, and that these features will co-travel with her broader metabolic, inflammatory, and thyroid status. Our findings caution against this assumption. Two women with similar endocrine phenotypes may have divergent metabolic risk, and two women with comparable metabolic profiles may differ substantially in reproductive endocrine severity or thyroid autoimmune status. A classification built from any single biological domain will therefore capture only part of a patient’s overall risk architecture.

A practical implication is that the biological space most relevant for clinical decision-making depends on the clinical question being asked. Endocrine phenotyping is likely most informative when the question concerns hyperandrogenism, menstrual irregularity, or fertility. Metabolic phenotyping is more relevant for cardiometabolic risk stratification and screening for diabetes and dyslipidemia. Inflammatory phenotyping may eventually help identify women who could benefit from interventions targeting chronic low-grade inflammation. Thyroid phenotyping, although exploratory in the present study, underscores that thyroid status should not be assumed to be predictable from reproductive or metabolic profile alone. This domain-specific approach is closer in spirit to precision medicine than a search for a single unifying PCOS phenotype, because it tailors the choice of biological information to the decision actually being made, rather than forcing every clinical decision through one classification.

Although our findings are not intended to change current diagnostic criteria for PCOS, they suggest that comprehensive phenotyping spanning endocrine, metabolic, inflammatory, and thyroid domains, rather than reliance on any single one, may better support individualized risk assessment. Prospective studies linking cross-space phenotypes to treatment response and long-term outcomes will be needed to establish whether this approach improves clinical decision-making in practice.

### 4.5. Methodological implications

Beyond its biological findings, this study introduces a methodological framework for investigating disease heterogeneity across multiple physiological domains. Most previous unsupervised learning studies in PCOS have combined diverse clinical, endocrine, metabolic, and biochemical variables into a single feature space before clustering. Although this strategy aims to capture overall disease heterogeneity, it implicitly assumes that all measured variables reflect a common latent biological structure.

Our findings challenge this assumption. By independently deriving phenotypes within predefined biological spaces and subsequently quantifying agreement between them, we demonstrate that different physiological domains can produce highly stable yet only partially concordant clustering solutions. These observations suggest that disagreement between clustering studies may not necessarily indicate methodological inconsistency but may instead arise because different feature sets capture different aspects of disease biology.

The cross-space phenotyping framework therefore provides an additional layer of model evaluation that is largely absent from current phenotype discovery studies. Conventional clustering analyses typically assess internal performance using metrics such as silhouette score, information criteria, or cluster stability. While these measures are essential for evaluating the quality of individual clustering solutions, they do not address whether phenotypes identified from different biological domains represent the same underlying patient structure. Quantifying cross-space agreement directly provides complementary information regarding the biological consistency and interpretability of independently derived phenotypes.

Another important methodological aspect of the present study is the use of independent non-circular biological validation. Phenotypes were evaluated using variables that were not involved in cluster derivation, thereby reducing the risk of circular interpretation. This distinction is particularly relevant for machine learning applications in biomedicine, where clustering results are frequently interpreted using the same variables that generated the clusters, potentially leading to overestimation of biological relevance.

Finally, our framework is not specific to PCOS. The concept of cross-space phenotyping may be applicable to other heterogeneous disorders characterized by interacting physiological systems, including diabetes, metabolic dysfunction-associated steatotic liver disease, autoimmune diseases, cardiovascular disorders, and neuropsychiatric conditions. More broadly, our findings illustrate that evaluating agreement between independently derived biological representations may provide important complementary information beyond conventional measures of clustering performance and may improve the interpretation of unsupervised learning in biomedical research.

### 4.6. Strengths and limitations

This study has several important strengths. First, to our knowledge, it is the first investigation to systematically compare independently derived biological phenotypes across multiple physiological domains within the same well-characterized PCOS cohort. Rather than identifying a single clustering solution, we explicitly evaluated whether endocrine, metabolic, inflammatory, and thyroid phenotypes described the same underlying patient structure.

Second, all clustering analyses were performed using a predefined analytical pipeline that incorporated standardized preprocessing, dimensionality reduction, objective model selection, bootstrap stability assessment, and independent non-circular biological validation. This comprehensive framework minimized subjective decisions during phenotype discovery and strengthened the robustness and interpretability of the identified biological subgroups.

Third, phenotype robustness was evaluated using multiple complementary approaches. In addition to internal cluster stability, we assessed cross-space agreement, independent biological validation using variables excluded from phenotype derivation, and sensitivity analyses comparing complete-case and imputed datasets. Together, these analyses consistently supported the reproducibility and biological coherence of the identified phenotypes.

Several limitations should also be considered. First, this was a cross-sectional study, precluding assessment of temporal changes in phenotype membership or causal relationships between the investigated biological domains. Longitudinal studies are required to determine whether cross-space phenotypes remain stable over time and whether they predict clinically relevant outcomes.

Second, the study was conducted in a single tertiary referral center, which may limit the generalizability of the identified phenotype structures to other populations with different demographic, ethnic, or clinical characteristics. Independent external validation in geographically and ethnically diverse cohorts will therefore be essential.

Third, although the thyroid phenotypes demonstrated excellent numerical stability, one identified subgroup comprised a relatively small proportion of the study population. While this solution satisfied the predefined model-selection criteria, the biological interpretation of thyroid phenotypes should be considered exploratory until confirmed in larger independent datasets.

Fourth, biological spaces were defined a priori based on current biological knowledge rather than learned directly from the data. This choice was deliberate, as it allowed cross-space agreement to be interpreted in relation to well-established physiological domains; however, it also means that our findings cannot exclude the possibility that a data-driven grouping of variables would identify a different, or more finely resolved, partitioning of biological space. Future work using purely data-driven approaches to define candidate biological spaces would provide a useful complement to the present, knowledge-based framework.

Finally, the present study focused on four predefined biological spaces selected on the basis of clinical relevance and data availability. Other physiological domains, including genetic, transcriptomic, proteomic, metabolomic, microbiome, or imaging-derived features, were not available for analysis and may provide additional complementary dimensions of PCOS heterogeneity.

Despite these limitations, the consistency of findings across bootstrap resampling, cross-space agreement analyses, independent biological validation, and sensitivity analyses supports the robustness of our principal conclusion that PCOS heterogeneity cannot be adequately represented by a single biological phenotype.

### 4.7. Conclusion

Using a cross-space phenotyping framework, we show that independently derived endocrine, metabolic, inflammatory, and thyroid phenotypes in women with PCOS are each reproducible and biologically valid, yet only partially concordant with one another.

These findings indicate that PCOS heterogeneity is intrinsically multidimensional, and offer a parsimonious explanation for why more than a decade of PCOS clustering studies has failed to converge on a single phenotype structure: no single structure exists for the literature to converge on. Different biological spaces, and the specific variable combinations used to define them, will continue to yield different, individually valid, partitions of the same population.

Beyond PCOS, we offer cross-space phenotyping as a general strategy for other heterogeneous diseases in which multiple physiological systems interact only partially. Prospective, longitudinal studies incorporating molecular data and clinical outcomes will be needed to determine whether this framework improves patient stratification and supports precision medicine approaches in PCOS.

## Data Availability

The clinical dataset analyzed during the current study is not publicly available because it contains sensitive patient information and institutional ethical restrictions prohibit unrestricted public release. De-identified data may be made available from the corresponding author upon reasonable request and subject to approval by the appropriate institutional and ethical authorities.

https://github.com/npiorkowska-science/pcos-cross-space-phenotyping

## Code Availability

The complete analytical workflow developed for this study, including Python scripts for data harmonization, preprocessing, missing-data handling, dimensionality reduction, clustering, stability assessment, cross-space agreement analysis, biological validation, sensitivity analyses, and figure generation, is openly available in the following GitHub repository: https://github.com/npiorkowska-science/pcos-cross-space-phenotyping

The repository also contains documentation describing the computational workflow, software dependencies, and instructions required to reproduce all analyses presented in this manuscript.

## Ethics Approval

This study was conducted in accordance with the Declaration of Helsinki and was approved by the Bioethics Committee of Wroclaw Medical University (approval no. 254/2021).

## Consent to Participate

Written informed consent was obtained from all participants prior to inclusion in the study.

## Consent for Publication

All authors have reviewed and approved the final version of the manuscript and consent to its publication.

## Author Contributions

Natalia Piórkowska: Conceptualization, methodology, software, formal analysis, investigation, data curation, visualization, writing – original draft, writing – review and editing.

Alan Ostromęcki: Methodology, software validation, formal validation, writing – review and editing.

Grzegorz Franik: Data curation.

Anna Bizoń: Writing – review and editing, supervision.

All authors read and approved the final manuscript.

## Acknowledgements

The authors thank all study participants and the clinical staff involved in patient recruitment, data collection, and laboratory diagnostics.

## Supplementary Materials

### Supplementary Table

Supplementary Table S1. Complete biological characterization of endocrine, metabolic, inflammatory, and thyroid phenotypes.

Supplementary Table S2.

